# Development of a Deep-Learning Algorithm for Detecting Suspicious Breast Lesions on Chest CT

**DOI:** 10.1101/2025.01.24.25321095

**Authors:** Hongyi Zhang, Jacob Johnson, Lawrence Ngo

**Affiliations:** University of Texas Southwestern School of Medicine, Dallas, TX; Covera Health, New York, NY; Duke Department of Radiology, Duke University School of Medicine, Durham, NC

## Abstract

A convolutional neural network (CNN) was trained and evaluated for detecting suspicious breast lesions on a large dataset of chest CT exams from a teleradiology practice covering over 2,000 hospital sites. Radiologists annotated any discrete nodules or masses appearing within breast tissue, and the model was tested on a held-out set. At a threshold achieving 0.99 specificity, the model demonstrated a sensitivity of 0.32 and a positive predictive value (PPV) of 0.50. In a scenario where sensitivity and specificity were balanced, the model achieved sensitivity and specificity of 0.79 each, resulting in a PPV of 0.10. The overall performance, as indicated by the area under the receiver operating characteristic curve (AUC), was 0.87. These results highlight the potential of an automated system to identify suspicious breast lesions on chest CT exams, thereby aiding in the opportunistic detection of malignancies.

## Introduction

Breast cancer remains one of the most common causes of cancer mortality among women worldwide, with more than two million new cases each year (1,2). While dedicated mammography and other breast imaging modalities constitute the primary screening approaches, a vast number of chest computed tomography (CT) studies are obtained daily for non-breast indications. It is widely recognized that these examinations can incidentally reveal suspicious breast lesions (3,4). However, because the primary focus of chest CT interpretations typically lies in the lungs, mediastinum, or cardiovascular structures, subtle breast nodules or masses may be missed.

The present work centers on a large, multi-site teleradiology practice that reads chest CTs for over 2,000 hospital sites. The scans depict portions of breast tissue, offering an opportunity for opportunistic lesion detection. Motivated by recent advances in machine learning–based medical image analysis, a convolutional neural network (CNN) was developed to localize suspicious focal findings in the breast region. The objective was to determine if such a model could reliably identify nodules or masses that otherwise might go unnoticed, thereby improving the rate at which clinically significant breast lesions are flagged for further evaluation.

Despite encouraging early results in computer-assisted detection, challenges persist. Chest CT protocols are not optimized for breast imaging, the degrees of breast compression and contrast enhancement vary widely, and breast anatomy may appear only partially in the field of view.

Moreover, breast implants and CT artifacts can mimic focal lesions. Nevertheless, a reasonably accurate automated tool could greatly reduce the likelihood of missed malignancies.

Below, we describe our data collection, modeling strategy, and results from a retrospective evaluation. We demonstrate performance at two different operating thresholds—one focused on high specificity, the other on a more balanced tradeoff. Finally, we note current limitations of this study and highlight directions for prospective validation, which would be of interest to any reviewer seeking to see this approach fully integrated into clinical practice.

## Methods

### Data Collection and Annotations

We obtained a retrospective dataset of chest CT scans performed between 2017 and 2020 at a major teleradiology practice reading for more than 2,000 hospital sites. Both contrast-enhanced and non-contrast exams were included, provided they contained at least partial depiction of breast tissue. All data were fully de-identified before analysis.

Board-certified radiologists used RectLabel, an imaging labeling tool, to place bounding boxes around any discrete nodules or masses in the breast region. The tool enabled annotations, facilitating the identification and demarcation of spiculated or lobulated shapes that stood apart from the background. Findings clearly benign in nature (e.g., characteristic lymph nodes, simple cysts) were excluded from the ground-truth annotations.

### Model Architecture and Training

We employed a “2.5D” detection approach to leverage limited axial context while retaining a standard 2D bounding-box framework. Specifically, the input consisted of several contiguous slices stacked as separate channels. For the detection engine, we used the implementation of a YOLO-style architecture from Ultralytics (https://github.com/ultralytics/yolov5). The model output was a set of bounding boxes with associated confidence scores for suspicious lesions.

Training and validation sets were split so that no patient appeared in both. The training set consisted of 14,105 positive samples and 26,195 negative images. Data augmentations (random flips, slight rotations, and intensity shifts) were applied to enhance robustness. After tuning hyperparameters (e.g., batch size, learning rate) on the validation set, the final model checkpoint was tested on a held-out cohort. We derived per-exam sensitivity, specificity, and various other metrics at multiple confidence-score thresholds.

## Results

In the test set (104 positive and 3,474 negative), the model achieved an overall area under the receiver operating characteristic curve (AUC) of 0.87 (Figure 1). We then selected two decision thresholds:

**Figure 1.**
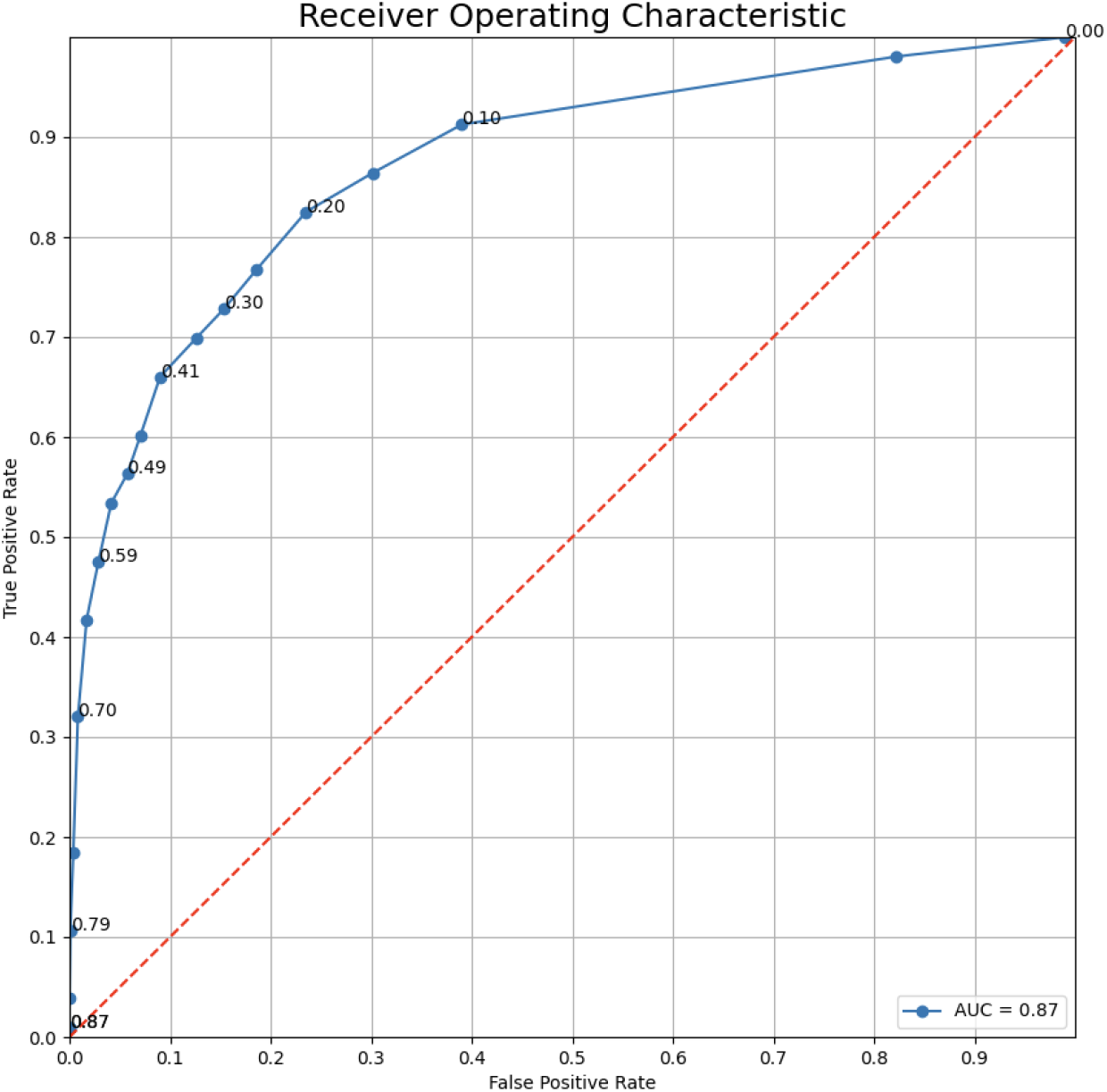
Receiver Operating Characteristic (ROC) curve for the performance of the breast lesion algorithm at varying thresholds.

**Figure 2.**
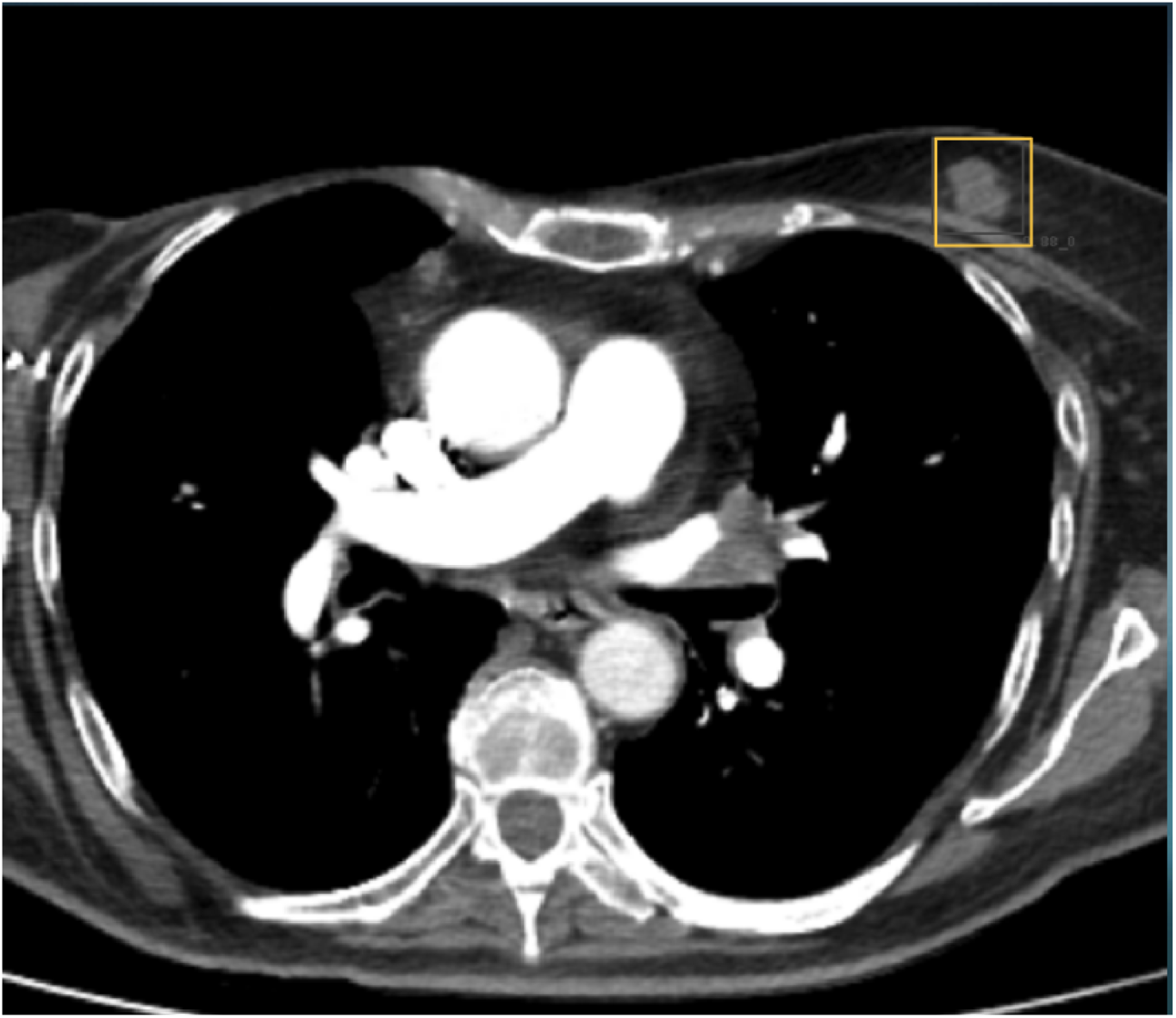
True positive detection of a suspicious breast lesion in the left breast.

1. High-Specificity Threshold: With specificity set to 0.99, the sensitivity was 0.32 and the positive predictive value (PPV) was 0.50.
2. Balanced Threshold: With a relaxed threshold providing sensitivity of 0.79 and specificity of 0.79, the PPV was 0.10 (Table 1, Figure 3).

**Table 1.** Performance of the model at two operating thresholds: High-Specificity and Balanced.

| Threshold | Sensitivity | Specificity | PPV |
| --- | --- | --- | --- |
| High-Specificity | 0.32 | 0.99 | 0.50 |
| Balanced | 0.79 | 0.79 | 0.10 |

**Figure 3.**
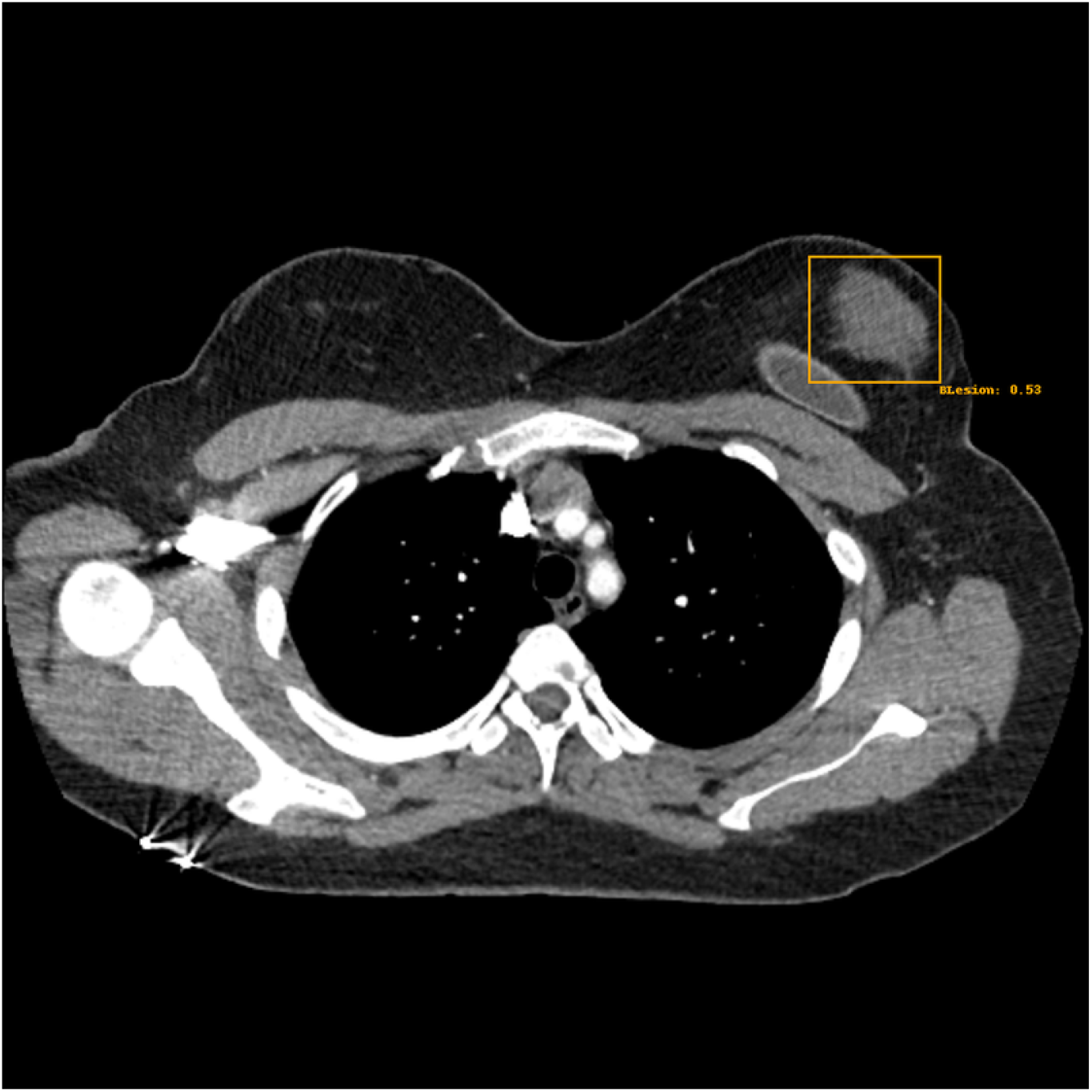
False positive detection on an axial CT of normal parenchymal tissue adjacent to a left breast implant.

In practice, therefore, if one wishes to capture a greater fraction of true lesions (sensitivity ~0.79) while accepting moderate false positives (specificity ~0.79), only about 1 in 10 flagged findings would be genuine suspicious lesions at this threshold.False positives were commonly caused by the edges of breast implants (Figure 3), while a smaller portion stemmed from dense fibroglandular tissue mischaracterized as nodular masses. Enhancing training with more negative examples of implants and a broader spectrum of benign tissue appearances could help mitigate these errors.

## Discussion

This study demonstrates that a single CNN-based approach, incorporating 2.5D inputs and an object detection backbone, can effectively identify focal breast nodules or masses on chest CT scans from a large, diverse teleradiology dataset. The model’s performance, with an AUC of 0.87 across the test set, suggests its potential utility; however, optimal threshold selection depends on clinical context. A high-specificity threshold may be preferred in settings prioritizing minimal false alarms, whereas a balanced threshold could be used where capturing most lesions is essential, even at the expense of more false positives. Regardless of the threshold, the system serves as an automated safety-net tool, flagging potential breast lesions in scans primarily obtained for non-breast indications such as thoracic or cardiac evaluations.

Although the performance metrics are encouraging, prospective clinical validation is needed to determine if flagged lesions prompt additional imaging or earlier diagnoses, ultimately improving patient outcomes. Aligning with the broader recognition of the role of incidental findings, this tool underscores how breast abnormalities detected on CT can contribute to early cancer diagnoses. To further enhance the system, future refinements such as training with implant-specific negative examples or advanced morphological filters may help reduce false positives. Additional data representing a broader spectrum of benign breast conditions could also bolster model robustness. Comparing the model’s performance with that of human readers is another key step, as such evaluations would elucidate its utility both as a standalone tool and in augmenting radiologists.

In conclusion, this system demonstrates promising accuracy for opportunistic breast lesion detection on routine chest CT. As the volume of chest CT scans continues to rise, integrating computer-assisted detection systems like this could help radiologists identify crucial incidental findings, fine-tune workflows, and ultimately improve patient care.

## Data Availability

The data from this manuscript is proprietary and not publically available.

## Acknowledgements

We would like to acknowledge Nadine Ly for assistance in the coordination and execution of this project.

